# Associations between Traditional Chinese Medicine Body Constitution and Cardiovascular Disease Risk in a White population

**DOI:** 10.1101/2022.12.13.22283433

**Authors:** Lihua Shu, Xiaolin Yin, Xiangzhu Zhu, Jing Zhao, Xinqing Deng, Yevheniy Eugene Shubin, Harvey J. Murff, Reid M. Ness, Chang Yu, Martha J. Shrubsole, Qi Dai

## Abstract

**Introduction:** Traditional Chinese medicine (TCM) has guided generations of practice on disease treatment and health maintenance. The TCM principles include the framework of body constitution (BC). In essence, it represents one of the first attempts at applying the principle of personalized, precision medicine. Major limitations to broad implementation of the body constitution (BC) framework, and perhaps TCM as a whole, include not only a lack of empirical study about its relation to other models of health maintenance but also a poor understanding of its applicability outside of Chinese population.

**Methods:** We conducted a study using baseline data from the Personalized Prevention of Colorectal Cancer Trial. 191 participants from an almost entirely White population were evaluated for BC type.

**Results:** Fifty-seven (29.8%) were identified as neutral BC, while Blood-stasis (17.3%), Qi-deficient (13.6%), and Special diathesis (10.5%) were the pre-eminent pathologic subtypes. We also found there are substantial differences in proportions of TCM BC types in our study of white Americans conducted in US from previous studies conducted in Chinese populations. Additional analyses investigated the relationship between cardiovascular disease (CVD) risk and BC subtypes. Among them, Yang-deficiency, Yin-deficiency, and Blood stasis carried a lower risk of CVD.

**Conclusions:** It is important to understand the underlying mechanisms contributing to these differences, which may not only help to understand the underlying mechanism for TCM, but also help to identify novel factors or mechanisms for CVD risk, prevention and treatment.

## Introduction

In Traditional Chinese Medicine (TCM), a primary framework of health maintenance and surveillance hinges upon the classification of individuals into nine categories of “body constitutions” (BC): Gentleness type, Qi-deficiency type, Yang-deficiency type, Yin-deficiency type, Phlegm-dampness type, Dampness-heat type, Blood-stasis type, Qi-depression type, and Special diathesis type ^1^. These BCs help to draw a picture of an individual’s health status and have been used on a provider-specific basis to identify patients who may be at risk for future disease, such as cardiovascular disease (CVD). However, TCM has a very limited role in Western medical practice and part of the reason for this is a simple lack of data concerning its application in Western populations. Additionally, while BCs are generally assigned based on different measures of an individual’s health, there exists little in the way of standardization when it comes to broadly applying this health framework in the clinical setting ^2-5^. What is more, there have been few studies regarding the relationship between TCM principles and contemporary models of health and disease. For example, the BC types of Yin and Yang-deficiency, which describe individuals who are routinely hot/anxious or cold/sluggish, seem to hint at manifestations of thyroid disease. Despite these parallels, little has been done to study what possible relations may exist between the BC framework and our current understanding of health and disease. Studies of this kind may not only help us understand more about the BC principle, but may also establish a role for BCs in current medical practices, such as in diagnostic screens.

Su et al. developed the most commonly used questionnaire to stratify individuals into the 9 major BC types ^6,7^. With this more standard approach, a number of studies have attempted to then explore utility of the TCM BC structure, specifically with the intent to identify a biological basis for the different BC types. In one particular study, associations between TCM BC types and HLA polymorphisms were identified ^8^. In a similar study, it was discovered that specific BC types are more likely to carry genetic variations of metabolic genes ^9^. These efforts represent attempts to ground the principles of the TCM BC framework in empiric data that explores physiologic and genetic bases for the different TCM BCs. This will in turn help identify clinical associations to chronic disease.

CVD is of particular interest in this study. It remains the leading cause of death worldwide, in particular due to its subtypes of coronary artery disease (CAD), as well as its manifestations of acute coronary syndrome (ACS) and myocardial infarction (MI) ^10,11^. The landmark Framingham Heart study provided data to establish what are now well-known risk factors of CVD, including cigarette smoking, cholesterol, blood pressure, and others ^12^. Understanding and identifying these risk factors have led to major advances in health maintenance, including the advent of routine blood pressure monitoring, regular exercise, and lipid control. Nonetheless, heart disease remains the leading cause of death in the US. Thus, there is a need to identify other early markers of CVD or populations at greatest risk to further preventive cardiology. The relationship of TCM BCs and markers of chronic disease, including dyslipidemia, hypertension, diabetes, etc., have further been studied and helped foster increased interest in the TCM BC system, as they might serve as an early indicator of needed intervention for specific chronic illnesses ^13-15^.

However, all of these previous studies using TCM BC have been conducted in Chinese populations except for one study of 400 White college students attending three Beijing universities in China ^16^. In addition, none of the previous studies have examined the associations between TCM BCs and CVD risk. To that end, the aim of this study is to investigate the ability for TCM BCs to identify participants at higher risk for CVD per general cardiovascular risk score (GCRS), an updated version of the original Framingham risk score (FRS), in a US population of white individuals; whether TCM BC is a better correlate of biomarkers of CVD (C-reactive protein (CRP) and uric acid) ^17,18^ than the GCRS; and whether TCM BC is associated with weight, body mass index and waist-to-hip ratio. In the same way that the Framingham study gave physicians insight on which health parameters may predict later heart disease, we seek to determine the potential value of utilizing TCM BCs in the clinical setting to predict CVD development ^19,20^.

## Materials and Methods

We conducted a cross-sectional study using baseline data from the Personalized Prevention of Colorectal Cancer Trial (PPCCT; NCT01105169 at ClinicalTrials.gov). The primary aim of the PPCCT was to examine the effect of Mg supplementation on expression of carcinogenesis biomarkers in colorectal mucosa. Between 2010 and 2017, the PPCCT enrolled 250 participants, aged 40 to 85 years, with high risk of colorectal cancer from Vanderbilt University Medical Center. Detailed inclusion and exclusion criteria have been reported (Supplemental materials) ^21,22^. The study was approved by the Vanderbilt Institutional Review Board.

### Constitution in Traditional Chinese Medicine Questionnaire

A self-administered Traditional Chinese Medicine Questionnaire (TCMQ) was implemented for a subset of participants enrolled from May 31, 2012 to Jan 30, 2016 (n=191). The original version of the standardized Constitution in TCMQ was developed in China with sensitivity and specificity of predicting BC types ranging from 42.7% to 82.7% ^1,7^. The TCMQ was translated to English by the research team and tested in cognitive debriefing interviews before it was applied in the study. The TCMQ consists of 60 questions, which fall into nine subscales that correspond to one of nine TCM BC types: Gentleness (8 Items), Qi-deficiency (8 Items), Yang-deficiency (7 Items), Yin-deficiency (8 Items), Phlegm-wetness (8 Items), Wetness-heat (6 Items), Blood-stasis (7 Items), Qi-depression (7 Items), and Special diathesis (7 Items). Six of the questions exhibit overlap between different subscales.

The 60-item questionnaire is graded on a 5-point Likert scale, ranging from 1 (not at all) to 5 (very much). Each of the nine subscales within the TCMQ assess one type of the TCM BC individually. A total score of each subscale is obtained by summing relevant item scores ^23^. Then, the transformed score was generated for each type by using the following equation.

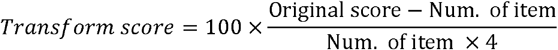

Following the criteria, a higher score in a specific TCMQ BC subscale indicates a higher likelihood of the corresponding BC type, with a score of 30 being a “threshold” for case definition. To ultimately classify a participant within a specific TCM BC, the following algorithm is applied: when 1) the score for the Gentleness subscale is greater than or equal to 60 and other BC type scores are less than 30, the study participant is diagnosed as “Gentleness” which is considered the neutral BC; when 2) the score for an imbalanced BC type (all other BCs) is greater than or equal to 40, then the participant is regarded as one or more of eight imbalanced BC types; when 3) a BC type score is between 30-40, the diagnosis will be made by a well-trained Chinese Medicine Practitioner. Based on the TCM theory, coexistence of multiple imbalanced BC types was possible.

### General cardiovascular risk score

Participants completed a telephone interviewer-administered survey at baseline to solicit information on medical history, tobacco and alcohol use, and other risk factors for colorectal cancer. Persons who smoked cigarettes regularly, measured by packs per day, during the previous 12 months were classified as smokers. During each clinic visit, the participant’s use of medications and biosamples were collected. Weight, height, waist and hip circumference as well as systolic and diastolic blood pressure, were measured. Body mass index (BMI) (kg/m^2^) and waist hip ratio (WHR) were calculated. Serum samples were assayed for a lipid profile (low-density lipoproteins cholesterol (LDL-C), high-density lipoproteins cholesterol (HDL-C), total cholesterol (TC), and triglycerides, c-reactive protein (CRP), and uric acid at the Vanderbilt Lipid Laboratory which is standardized by the Centers for Disease Control and Prevention for lipid analysis (Supplemental materials). Those used antihypertensive medication, blood pressure categorization was made accordingly. Participant characteristics were then assigned specific point values, with total points being then converted to a general cardiovascular risk score, representing a person’s 10yr risk of CVD. This score is a derivative of the original Framingham risk score (FRS) produced by the same group. However, it was reported to be better in predicting CVD risk, which may be explained by the modeling of risk factors continuous variables (as opposed to categories in the model developed by Wilson et al) ^24^. A risk assessment calculator based on the Cox regression model of proportional hazards was utilized to obtain participant GCRS ^25^.

Females:

Risk factors = (ln(Age) * 2.32888) + (ln(Total cholesterol) * 1.20904) - (ln(HDL cholesterol) * 0.70833) + (ln(Systolic BP) * Hypertension medication factor) + Cig + DM - 26.1931

Risk = 100 * (1 - 0.95012e(Risk factors))

Hypertension medication factor: 2.76157 (not on medication) vs 2.82263 (on medication) Cig = 0.52873

DM = 0.69154

Males:

Risk factors = (ln(Age) * 3.06117) + (ln(Total cholesterol) * 1.12370) - (ln(HDL cholesterol) * 0.93263) + (ln(Systolic BP) * Hypertension medication factor) + Cig + DM - 23.9802

Risk = 100 * (1 - 0.88936e(Risk factors))

Hypertension medication factor: 1.93303 (not on medication) vs 1.99881 (on medication) Cig = 0.65451

DM = 0.57367

HDL: high-density lipoproteins; BP: blood pressure; Cig: cigarette smoking status; DM: Diabetes.

### Statistical analyses

We performed generalized linear model for continuous variables or Pearson chi-squared tests for categorial variable to evaluate the differences among the nine types of TCM BC. We further evaluated the impact of significant differences among Gentleness group, Low CVD risk group and the remaining group. Mean ± standard deviation for continuous demographic variables and percentage for categorical demographic variables were presented in Table 2 and Table 3.

Additionally, we performed analyses stratified by sex. All P values are two sided and statistical significance was determined using an alpha level of 0.05. The data analyses used software SAS Enterprise Guide 7.1.

## Results

For the reliability and validity of the self-administered Traditional Chinese Medicine Questionnaire (English Version), the construct validity was confirmed with scaling success rates that ranged from 75.0 to 100% (Supplemental Table 2), and confirmatory factor analysis indicate an acceptable model fit. The results of reliability for test-retest reliability (intra-class correlation coefficients) were 0.7-0.8 (Supplemental Table 3), the Cronbach’s alphas ranged from 0.44 to 0.72 during three-month period.

As shown in Table 1, the distribution of BC types varied greatly between this study and previous studies conducted in Chinese populations. Gentleness (29.8%) was the predominant BC type in this study, while the three most common pathologic BC types were Blood-stasis (17.3%), Qi-deficient (13.6%), and Special diathesis (10.5%). In contrast, the most common pathologic BC types found in Chinese populations, in no particular order, were Qi-deficient, Yang-deficient, Yin-deficient, and Phlegm-Dampness.

**Table 1:** Comparative distribution of TCM constitution types among PPCCT population and Representative Study population in China.

| Characteristic | Study |  |  |  |  |  |  |
| --- | --- | --- | --- | --- | --- | --- | --- |
|  | PPCCT | Caucasian in China | Chinese Population |  |  |  |  |
|  |  | Jing et al. 2012 | Zhu et al. 2011 | Zhu et al. 2017 | Li et al. 2017 | Bai et al. 2021 | Jiang et al. 2018 |
| Sample size, n | 191 | 396 | 8448 | 2660 | 3748 | 313 | 708 |
| Age (y), Mean ± SD | 60 ± 7.8 | 34.1 ± 14.3 | 41.6 ± 15.9 | 52.5 ± 13.9 | 69.3 ± 7.6 | 77.1 ± 8.22<sup>1</sup> | 28.3 ± 3.0<sup>2</sup> |
| Gender, n (%) |  |  |  |  |  |  |  |
| *Female* | 91 (47.6) | 240 (60.6) | 4153 (49.2) | 1260 (47.4) | 2039 (54.4) | 268 (82.4) | 708 (100) |
| TCM Constitution type, % |  |  |  |  |  |  |  |
| *Gentleness* | 29.8 | 51.0 | 32.1 | 22.6 | 39.9 | 27.50 | 45.2 |
| *Qi-deficient* | 13.6 | 9.6 | 13.4 | 19.1 | 22.9 | 5.43 | 3.4 |
| *Yang-deficient* | 4.7 | 13.6 | 9.0 | 8.7 | 17.9 | 11.18 | 23.0 |
| *Yin-deficient* | 6.3 | 4.5 | 8.3 | 8.5 | 6.6 | 23.30 | 9.5 |
| *Phlegm-Dampness* | 8.4 | 5.1 | 7.3 | 11.1 | 6.0 | 16.90 | 4.9 |
| *Dampness-Heat* | 3.1 | 3.5 | 9.1 | 9.3 | 2.5 | 0.64 | 2.0 |
| *Blood-Stasis* | 17.3 | 2.5 | 8.1 | 8.1 | 1.5 | 3.83 | 3.5 |
| *Qi-depression* | 6.3 | 4.0 | 7.7 | 7.3 | 1.1 | 8.95 | 6.4 |
| *Special Diathesis* | 10.5 | 6.1 | 5.0 | 5.4 | 1.7 | 2.24 | 2.1 |

Continuous variables were presented as Mean ± SD; categorical variables were presented as n (%) or %
<sup>1</sup>: Study conducted among an elderly population in Macau
<sup>2</sup>: Study was done in a population of only women of childbearing age

The characteristics of the study participants are shown overall and by BC type in Table 2. Among the 191 participants, the median age was 60, a minority were active smokers, and a majority were using anti-hypertensive and anti-lipid medications, both of which reflect a higher risk of CVD. There were statistically significant differences in the distribution of several factors across the various BC types. These included BMI, gender, CRP, and GCRS. Specifically, the Gentleness BC had a generally lower average BMI (28.9), CRP (2.2), and GCRS (20.6), as well as lower female predominance (26.3%) compared to other groups.

**Table 2:** Baseline Characteristics of Participants Overall and Grouped by TCM BC Type.

|  |  |  | Qi- | Yang- | Yin- | Phlegm- | Dampness- |  |  | Special |  |
| --- | --- | --- | --- | --- | --- | --- | --- | --- | --- | --- | --- |
|  | Overall | Gentleness | deficiency | deficiency | deficiency | dampness | heat | Blood-stasis | Qi-depression | Diathesis |  |
| Characteristic | (n=191) | (n=57) | (n=26) | (n=9) | (n=12) | (n=16) | (n=6) | (n=33) | (n=12) | (n=20) | P value |
| Age (y), Mean ± SD | 60.0 ± 7.8 | 61.0 ± 6.9 | 62.6 ± 7.8 | 55.0 ± 4.2 | 58.7 ± 5.7 | 60.4 ± 9.7 | 61.5 ± 9.3 | 58.2 ± 8.1 | 59.3 ± 7.9 | 59.6 ± 9.8 | 0.28 |
| BMI (kg/m<sup>2</sup>), Mean ± SD | 30.3 ± 6.7 | 28.9 ± 6.7 | 32.4 ± 7.2 | 31.4 ± 7.1 | 32.9 ± 7.2 | 34.2 ± 6.3 | 30.5 ± 6.3 | 28.1 ± 4.0 | 31.9 ± 7.4 | 29.4 ± 7.1 | 0.03 |
| Gender, n (%) |  |  |  |  |  |  |  |  |  |  | <0.0001 |
| Male | 100 (52.4) | 42 (73.7) | 16 (61.5) | 1 (11.1) | 2 (16.7) | 6 (37.5) | 5 (83.3) | 8 (24.2) | 8 (66.7) | 12 (60) |  |
| Systolic Blood Pressure (mmHg), Mean ± SD | 127.5 ± 15.3 | 129.1 ± 13.2 | 128.5 ± 18.3 | 117.7 ± 14.7 | 120.0 ± 16.3 | 130.4 ± 10.9 | 134.2 ± 9.8 | 128.9 ± 17.8 | 120.7 ± 16.6 | 128.0 ± 13.7 | 0.16 |
| Total cholesterol (mg/dL), Mean ± SD | 204.7 ± 57.7 | 198.7 ± 54.7 | 198.1 ± 52.3 | 191.3 ± 37.3 | 216.8 ± 59.7 | 226.8 ± 70.9 | 162.2 ± 43.0 | 215.4 ± 69.9 | 201.1 ± 38.5 | 209.2 ± 55.8 | 0.36 |
| HDL (mg/dL), Mean ± SD | 61.2 ± 21.3 | 59.3 ± 20.8 | 58.8 ± 16.2 | 66.2 ± 9.3 | 61.0 ± 16.2 | 65.4 ± 36.2 | 44.7 ± 15.9 | 66.9 ± 23.3 | 58.3 ± 16.9 | 62.2 ± 18.5 | 0.42 |
| LDL (mg/dL), Mean ± SD | 118.5 ± 41.9 | 115.6 ± 43.2 | 112.9 ± 39.0 | 107.7 ± 28.9 | 126.3 ± 43.8 | 130.4 ± 40.5 | 96.5 ± 29.0 | 123.8 ± 49.3 | 120.8 ± 30.2 | 121.5 ± 44.8 | 0.72 |
| C-reactive Protein | 3.1 ± 3.9<br>(n=101) | 2.1 ± 2.1<br>(n=35) | 3.5 ± 3.6<br>(n=16) | 1.1 ± 1.1<br>(n=3) | 7.3 ± 4.8<br>(n=6) | 5.1 ± 5.8<br>(n=5) | 11.5 ± 15.5<br>(n=2) | 2.0 ± 1.2<br>(n=16) | 2.6 ± 3.7<br>(n=6) | 3.3 ± 3.4<br>(n=12) | 0.003 |
| Uric acid | 6.2 ± 1.7<br>(n=101) | 6.6 ± 1.8<br>(n=35) | 6.5 ± 2.0<br>(n=16) | 5.2 ± 1.5<br>(n=3) | 6.0 ± 1.4<br>(n=6) | 7.2 ± 1.3<br>(n=5) | 5.8 ± 0.8<br>(n=2) | 6.5 ± 0.8<br>(n=16) | 5.4 ± 1.3<br>(n=6) | 5.6 ± 1.3<br>(n=12) | 0.19 |
| General cardiovascular risk score | 18.2 ± 14.1 | 20.6 ± 14.2 | 21.9 ± 14.9 | 6.4 ± 3.8 | 13.2 ± 13.3 | 17.7 ± 12.5 | 27.4 ± 20.4 | 12.9 ± 10.4 | 20.0 ± 17.0 | 20.1 ± 14.7 | 0.01 |
| Smoking status, n (%) |  |  |  |  |  |  |  |  |  |  | 0.09 |
| Never | 102 (53.4) | 28 (49.1) | 16 (61.5) | 6 (66.7) | 6 (50) | 10 (62.5) | 4 (66.7) | 22 (66.7) | 4 (33.3) | 6 (30) |  |
| Ever/current | 89 (46.6) | 29 (50.9) | 10 (38.5) | 3 (33.3) | 6 (50) | 6 (37.5) | 2 (33.3) | 11 (33.3) | 8 (66.7) | 14 (70) |  |
| Drinking status, n (%) |  |  |  |  |  |  |  |  |  |  | 0.22 |
| Never | 70 (36.7) | 17 (29.8) | 13 (50) | 4 (44.4) | 6 (50) | 8 (50) | 2 (33.3) | 12 (36.4) | 5 (41.7) | 3 (15) |  |
| Ever | 42 (21.9) | 13 (22.8) | 4 (15.4) | 0 (0) | 4 (33.3) | 3 (18.8) | 3 (50) | 5 (15.2) | 4 (33.3) | 6 (30) |  |
| Current | 79 (41.4) | 27 (47.4) | 9 (34.6) | 5 (55.6) | 2 (16.7) | 5 (31.3) | 1 (16.7) | 16 (48.5) | 3 (25) | 11 (55) |  |
| Antihypertensive medication use, n (%) | 78 (40.84) | 21 (36.9) | 17 (65.4) | 2 (22.2) | 6 (50) | 6 (37.5) | 4 (66.7) | 12 (36.4) | 4 (33.3) | 6 (30) | 0.16 |
| Lipid control medication use, n (%) | 68 (35.6) | 19 (33.3) | 13 (50) | 2 (22.2) | 4 (33.3) | 3 (18.8) | 5 (83.3) | 12 (36.4) | 2 (16.7) | 8 (40) | 0.10 |
| Glycemic control medication use, n (%) | 22 (11.52) | 6 (10.6) | 5 (19.23) | 1 (11.1) | 2 (16.7) | 2 (12.8) | 2 (33.3) | 0 (0) | 1 (8.3) | 3 (15) | 0.31 |
| Physically active $\geq 2$ days/week, n (%) | 162 (84.8) | 52 (91.2) | 22 (84.6) | 6 (66.7) | 9 (75) | 12 (75) | 4 (66.7) | 28 (84.9) | 11 (91.7) | 18 (90) | 0.38 |
| Education college or beyond | 173 (90.6) | 53 (92.9) | 24 (92.3) | 9 (100) | 11 (91.7) | 12 (75) | 5 (83.3) | 32 (96.9) | 9 (75) | 18 (20) | 0.18 |

Continuous variables were presented as mean ± SD, *P* values were calculated using GLM model; categorical variables presented as n (%), *P* values were calculated using chi-square test.
LDL: low-density lipoproteins; HDL high-density lipoproteins

### Comparison of cardiovascular disease risk among TCM subtypes

Among participants using hypertensive medication, the three most common BC types were Gentleness (26.9%), Qi-deficient (21.8%), and Blood-stasis (15.4%) (results not shown in table). On the other hand, the predominant types among participants using lipid control medications were Gentleness (27.9%), Qi-deficient (19.1%), Blood stasis (17.7%), and Special diathesis (11.8%) (results not shown in table). The GCRS averages, which represent a 10-year risk of cardiovascular disease, were statistically significantly different across BC types (p=0.013). Specifically, individuals in Yang-deficiency (6.4), Blood stasis (12.9), and Yin-deficiency (13.2), had a substantially lower risk of cardiovascular disease than other groups (Table 2). These BC types were also more common among women than among men.

To further investigate the characteristics of the three BC types with lower CVD risk, we combined individuals of these BC types into one low-risk group, and compared this new group against both the normal “Gentleness” type and the combination of the other 5 TCM groups (Table 3). As previously noted, the low-risk group had a significantly higher proportion of women. Additionally, individuals in these groups were younger (57.8 y, p=0.047) than their counterparts in the rest of the study (61.0 y and 60.8 y, respectively). They also tended to have a higher average serum HDL-C (p=0.228), which is known to be protective against CVD, compared to counterparts. This low-risk group also had the highest percentage of never-smokers and never-drinkers compared to the other groups, although this difference was not statistically significant. In stratified analysis by sex, there were no statistically significant differences for BMI, CRP, uric acid, or the GCRS (Table 4) by BC type.

**Table 3:** Stratified analysis of neutral BC group, low CVD risk group, and remaining groups.

| Characteristics | Gentleness<br>(n=57) | Low CVD risk group<br>(n=54)<sup>1</sup> | Remaining groups<br>(n=80)<sup>2</sup> | P-value |
| --- | --- | --- | --- | --- |
| Age (y), Mean ± SD | 61.0 ± 6.9 | 57.8 ± 7.1 | 60.8 ± 8.7 | 0.047 |
| BMI (kg/m<sup>2</sup>), Mean ± SD | 28.9 ± 6.7 | 29.7 ± 5.7 | 31.7 ± 7.0 | 0.037 |
| Gender (n), % |  |  |  | <0.0001 |
| *Male* | 42 (73.7) | 11 (20.4) | 47 (58.8) |  |
| *Female* | 15 (26.3) | 43 (79.6) | 33 (42.3) |  |
| Systolic Blood Pressure (mmHg), |  |  |  |  |
| Mean ± SD | 129.1 ± 13.2 | 125.1 ± 17.4 | 128 ± 15.2 | 0.368 |
| Total cholesterol (mg/dL), Mean ± SD | 198.7 ± 54.7 | 211.7 ± 63.2 | 204.4 ± 56.1 | 0.498 |
| HDL (mg/dL), Mean ± SD | 59.3 ± 20.8 | 65.4 ± 20.0 | 59.8 ± 22.3 | 0.228 |
| LDL (mg/dL), Mean ± SD | 115.6 ± 43.2 | 121.7 ± 45.1 | 118.5 ± 39.2 | 0.749 |
| C-reactive Protein (mg/L), Mean ± SD | 2.1 ± 2.1 | 3.5 ± 4.3 | 3.8 ± 4.6 | 0.146 |
| CRP level (elevated), n (%) | 0 (0) | 3 (12) | 4 (9.8) | 0.128 |
|  | 6.6 ± 1.8 | 5.5 ± 1.3 | 6.3 ± 1.6 | 0.039 |
| Uric acid level (elevated), n (%) | 23 (65.7) | 7 (28) | 21 (51.2) | 0.0157 |
| General cardiovascular risk score | 20.6 ± 14.2 | 11.9 ± 10.5 | 20.7 ± 15.0 | 0.0005 |
| Smoking (n), % |  |  |  | 0.344 |
| *Never* | 28 (49.1) | 34 (62.9) | 40 (50.0) |  |
| *Ever* | 25 (43.9) | 17 (31.5) | 30 (37.5) |  |
| *Current* | 4 (7.02) | 3 (5.6) | 10 (12.5) |  |
| Drinking (n), % |  |  |  | 0.536 |
| *Never* | 17 (29.8) | 22 (40.7) | 31 (38.8) |  |
| *Ever* | 13 (22.8) | 9 (16.7) | 20 (25) |  |
| *Current* | 27 (47.4) | 23 (42.6) | 29 (36.3) |  |
| Antihypertensive medications, % | 21 (36.8) | 20 (37.0) | 37 (46.3) | 0.434 |
| Lipid control medications, % | 19 (33.3) | 18 (33.3) | 31 (38.8) | 0.742 |
| Glycemic control medications, % | 6 (10.6) | 3 (5.6) | 13 (16.3) | 0.158 |

<sup>1</sup> Consists of pooled participants from Yang-deficient, Yin-deficient, and Blood-Stasis BC subtypes
<sup>2</sup> Consists of pooled participants from Qi-deficient, Phlegm-Dampness, Dampness-Heat, Qi-depression, and Special diathesis BC subtypes.
CRP elevated:>3.0mg/L, uric acid elevated:>7.0 mg/dL
P values were calculated using GLM model; categorical variables presented as n (%), P values were calculated using chi-square test.

**Table 4:**
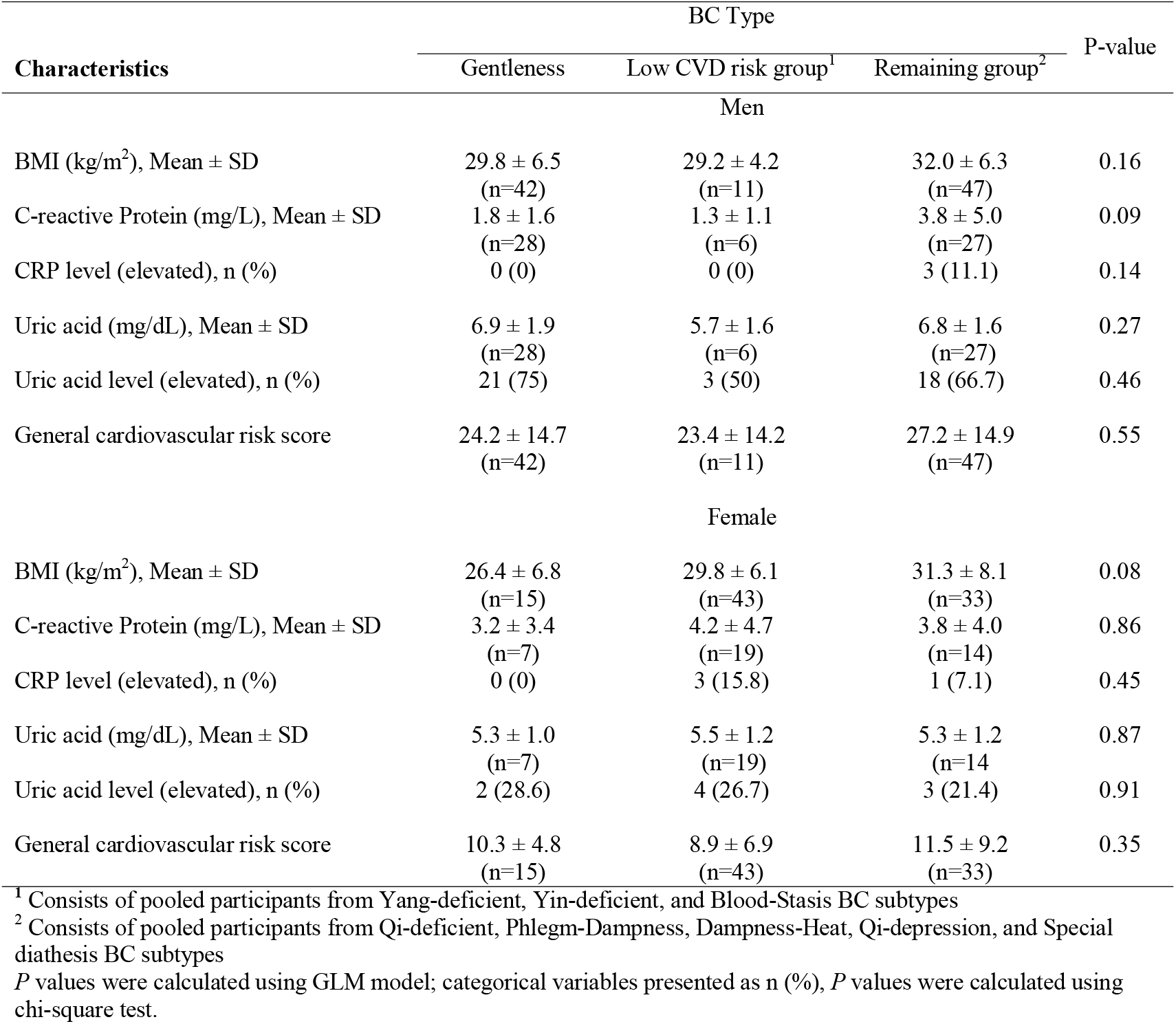
Analysis of neutral BC group, low CVD risk group, and remaining groups by sex.

### Stratified analyses by CRP and Uric acid levels

Based on previous studies which linked elevated CRP and uric acid to increased CVD risk, we classified our study population into “normal” and “elevated” groups. Specifically, individuals with CRP levels >10 mg/dL and uric acid >5.5 mg/dL were sorted into elevated CRP and uric acid groups, respectively. Average GCRS were 14.7 and 19.9 in the elevated and normal CRP groups, respectively; on the other hand, average GCRS was 21.7 and 16.8 in the elevated and normal uric acid groups, respectively. Analyses were then conducted to identify if specific TCM subtypes were associated with elevated biomarker levels. While there were no noticeable differences in the distribution of individuals with elevated CRP levels, the low-risk CVD group had a significantly lower level of elevated uric acid (Table 3). Further analysis of these findings occurred by stratifying the population by gender. Results of these analyses were not significant in terms of the distribution of individuals with elevated CRP/Uric acid levels (Table 4).

## Discussion

All previous studies of BC type have been conducted in Chinese populations except for a study of White college students in China ^16^. The present study, to the best of our knowledge, is the first to be done in an American population ^26^. The major types of TCM BC in our study of white individuals in the US were different from the types among Chinese in China. Furthermore, in this cross-sectional study, we newly examined the relationships between the nine BC types with predictive models or markers of CVD, in particular the GCRS.

Our study of white individuals in the US had a large proportion of Blood-Stasis (17.3%) and Special Diathesis types (10.5%). No previous study has been conducted in US populations. In a previous large study with 8,448 participants conducted within Chinese population, the most common pathologic BC subtypes are Qi-deficient (13.4%), Dampness-Heat (9.1%) and Yang-deficient (9.0%) ^27^. The difference between our study of white individuals in the US and this large study conducted in Chinese population may be due to multiple factors. First, our study participants may not represent the general US population. Thus, our finding needs to be confirmed by future larger studies conducted in US population. Secondly, this difference could be due to different genetic background. Thirdly, the difference could be caused by regional environmental factors as our study was conducted in one state in the US. There was one previous study of Caucasian college students attending three Beijing Universities in China ^16^. We found the distribution of major types of TCM BC in this earlier study is also different from that in our study. However, the White college students recruited in this earlier study were mostly younger than 50 years old and lived in Beijing, a Northern city in China while most of the participants in our study were older than 50 years and lived in a single US state. Thus, the differences could be caused by age and regional environmental factors.

Another potential explanation for these discrepancies may lie in identifying how biomarkers of common diseases contribute to an individual’s TCM subtype. In our study, significant differences were found among the nine different BC types by various different biomarkers and vitals for cardiovascular disease. Some of these findings were expected, such as lower BMIs in the “healthy” Gentleness controls and the classically thinner Blood-stasis compositions ^28^. However, the elevated BMIs in most other BC types, including the Phlegm-dampness and Yin-deficient types stand in contrast to findings in studies conducted in Chinese populations ^29^. Again, the reason is unclear. For example, White, and in particular White Americans, peoples tend to have a higher overall BMI than Asian populations ^30^. If BMI contributes to a person’s TCM BC type, then it may help to explain the differences in proportion of TCM BC subtypes between an American and Chinese populations. Taken a step further this could reflect differences in disease prevalence, such as cardiovascular disease, from one population to another, as previous studies found the incidence rates of cardiovascular disease and cancers (i.e. breast cancer, colorectal cancer and prostate cancer) are much higher in US populations than in Chinese populations ^31,32^. Zhu et al. also identified which TCM subtypes were more represented among major chronic diseases, including hypertension, hyperlipidemia, and hyperglycemia. Thus, further understanding the underlying mechanism for the difference in TCM BC type between US and Chinese population may help to identify the novel risk factors contributing to common disease etiology.

Another primary research focus of this study was the GCRS, which represents an individual’s percent risk of cardiovascular disease (CVD) in 10 years, with a higher score necessarily translating to a higher percent risk. Again, our analyses revealed significant differences in the overall GCRS across BC types. Constitutions with the lowest average GCRS included Yang deficient (6.4), Blood-stasis (12.9), and Yin deficient (13.2), while Dampness-Heat (DH) (27.4) and Qi-deficient (21.9) recorded the highest GCRS.

Further subgroup analysis of the three low-CVD risk groups (Yin/Yang-deficient and BS) revealed a significantly lower average age and BMI, as well as a higher predominance of women, compared to both the Gentleness group and the remaining 5 groups. This biologically makes sense in the context of the Framingham model, which typically associated men, particularly older and more obese men, with increased CVD risk. Another finding was a significantly lower uric acid level among the low-CVD risk group. While uric acid was not a component of the initial Framingham study, more recent literature has reported on an association between uric acid and CVD risk, specifically an association between elevated uric acid (>5.5) and increased CVD risk ^33^. Stratified analyses by sex revealed a loss of significance in the differences in BMI, GCRS, and Uric acid. Mediation analysis was also conducted, which revealed no significant effect mediation by sex. Thus, the loss of statistical significance when stratified by sex may be due to reduced sample size as we also found the patterns still remained in the stratified analysis.

The major limitation of the study was the sample size, specifically the sample sizes in each BC group, which limits the power of the analysis in stratified analysis. Secondly, the limitation is that not all parts of the survey were easily translatable to English from the original Chinese version due to the cultural differences, with some questions, such as those involving “sighing without reason” or “thick saliva” perhaps being less readily answered by our exclusively American population. However, this only appeared in limited survey questions which may not materially affect the directions of results. Also, this non-differential misclassification caused by the translation in cultural difference usually biases the results to the null. In other words, the true associations may be stronger than the ones we observed. Thirdly, the limitation is that we did not collect family history of cardiovascular disease and cannot adjust for this covariate in the current study. Further studies are warranted to examine whether family history of CVD contributes to the TCM BC types related to CVD risk. Finally, although the intra-class correlation coefficients ranging from 0.7 to 0.8 and the Cronbach’s alphas ranging from 0.44 to 0.72 are reasonable for reliability, these findings indicate that TCM BC types could vary during the three-month period in a study of White individuals in the US. Further studies are warranted to identify the factors contributing to the changes.

The majority of previous studies conducted in Mainland China which have investigated a similar topic have noted significant associations between CAD and particular TCM BC types, in particular Yin/Yang deficiency as well as Phlegm-dampness ^14,34^. These findings have helped practitioners in China to better identify patients at risk for CVD and therefore initiate a preventive health regimen. Additional studies have also postulated associations between BC types and chemokine receptor/ligan expression, which would hold implications for a whole host of pathologies, including immunology and oncology. Translating these promising findings to an American population could be beneficial, given the prevalence of CVD in the United States.

However, our findings found although there are similarities, there are substantial differences between these two populations. Future studies should aim to understand the underlying mechanisms contributing to these differences, which may not only help to understand the underlying mechanism for TCM, but also help to identify novel factors or mechanisms for CVD risk, prevention and treatment.

In conclusion, future larger studies in US populations, particularly population-based studies including multiple racial/ethnic groups are warranted to confirm our findings. Further, future studies are also needed to examine whether combined use of BC type with the Framingham risk score can improve the Framingham risk score in predicting future risk of CAD in clinical settings.

## Supporting information

Supplemental Marterial

## Data Availability

The datasets generated during and/or analyzed during the current study are available from the corresponding author on reasonable request.

## Abbreviations

ACS: acute coronary syndrome
BC: body constitutions
BMI: Body mass index
CAD: coronary artery disease
CVD: cardiovascular disease
CRP: c-reactive protein
eGFR: estimated glomerular filtration rate
GCRS: General cardiovascular risk score
HDL-C: high-density lipoproteins cholesterol
LDL-C: low-density lipoproteins cholesterol
MDRD: Modification of Diet in Renal Disease
MI: myocardial infarction
TC: total cholesterol
TCM: Traditional Chinese Medicine
WHR: waist hip ratio

## Acknowledgments

The authors are grateful to the participants for participating in PPCCT. Authors thank the study staffs for their participation.

## Author contributions

Qi Dai: contributed to the hypothesis development; Qi Dai, Martha Shrubsole, Xiaolin Yin, Xiangzhu Zhu: contributed to the study design; Qi Dai, Martha Shrubsole, Xiaolin Yin, Xiangzhu Zhu, Xinqing Deng and Yevheniy Eugene Shubin: conducted the research; Lihua Shu, Xiangzhu Zhu, Jing Zhao: performed statistical analysis; Lihua Shu, Xiangzhu Zhu, Qi Dai, Martha Shrubsole drafted the manuscript; All the authors: contributed to the data interpretation and manuscript revision and read and approved the final manuscript.

## References

1. Wong W, Lam CL, Wong VT, et al. Validation of the constitution in chinese medicine questionnaire: does the traditional chinese medicine concept of body constitution exist? Evid Based Complement Alternat Med 2013;2013(481491, doi:10.1155/2013/481491

2. O’Brien KA, Abbas E, Zhang J, et al. An investigation into the reliability of Chinese medicine diagnosis according to Eight Guiding Principles and Zang-Fu Theory in Australians with hypercholesterolemia. J Altern Complement Med 2009;15(3):259–66, doi:10.1089/acm.2008.02043.

3. O’Brien KA, Abbas E, Zhang J, et al. Understanding the reliability of diagnostic variables in a Chinese Medicine examination. J Altern Complement Med 2009;15(7):727–34, doi:10.1089/acm.2008.0554

4. Mist S, Ritenbaugh C, Aickin M. Effects of questionnaire-based diagnosis and training on inter-rater reliability among practitioners of traditional Chinese medicine. J Altern Complement Med 2009;15(7):703–9, doi:10.1089/acm.2008.0488

5. Birkeflet O, Laake P, Vollestad N. Low inter-rater reliability in traditional Chinese medicine for female infertility. Acupunct Med 2011;29(1):51–7, doi:10.1136/aim.2010.003186

6. Chen LL, Lin JS, Lin JD, et al. BCQ+: a body constitution questionnaire to assess Yang-Xu. Part II: Evaluation of reliability and validity. Forsch Komplementmed 2009;16(1):20–7, doi:10.1159/000197770

7. Su YC, Chen LL, Lin JD, et al. BCQ+: a body constitution questionnaire to assess Yang-Xu. Part I: establishment of a first final version through a Delphi process. Forsch Komplementmed 2008;15(6):327–34, doi:10.1159/000175938

8. Chen S, Lv F, Gao J, et al. HLA class II polymorphisms associated with the physiologic characteristics defined by Traditional Chinese Medicine: linking modern genetics with an ancient medicine. J Altern Complement Med 2007;13(2):231–9, doi:10.1089/acm.2006.6126

9. Wu Y, Cun Y, Dong J, et al. Polymorphisms in PPARD, PPARG and APM1 associated with four types of traditional Chinese medicine constitutions. J Genet Genomics 2010;37(6):371–9, doi:10.1016/S1673-8527(09)60055-2

10. Roger VL. Epidemiology of myocardial infarction. Med Clin North Am 2007;91(4):537-52; ix, doi:10.1016/j.mcna.2007.03.007

11. Virani SS, Alonso A, Aparicio HJ, et al. Heart Disease and Stroke Statistics-2021 Update: A Report From the American Heart Association. Circulation 2021;143(8):e254–e743, doi:10.1161/CIR.0000000000000950

12. Dawber TR, Kannel WB, Revotskie N, et al. Some factors associated with the development of coronary heart disease: six years’ follow-up experience in the Framingham study. Am J Public Health Nations Health 1959;49(1349–56, doi:10.2105/ajph.49.10.1349

13. Wu HK, Ko YS, Lin YS, et al. The correlation between pulse diagnosis and constitution identification in traditional Chinese medicine. Complement Ther Med 2017;30(107–112, doi:10.1016/j.ctim.2016.12.005

14. Huang YC, Lin CJ, Cheng SM, et al. Using Chinese Body Constitution Concepts and Measurable Variables for Assessing Risk of Coronary Artery Disease. Evid Based Complement Alternat Med 2019;2019(8218013, doi:10.1155/2019/8218013

15. Zhu YJ, Zhang HB, Liu LR, et al. Yin-Cold or Yang-Heat Syndrome Type of Traditional Chinese Medicine Was Associated with the Epidermal Growth Factor Receptor Gene Status in Non-Small Cell Lung Cancer Patients: Confirmation of a TCM Concept. Evid Based Complement Alternat Med 2017;2017(7063859, doi:10.1155/2017/7063859

16. Ming-hua B. Applying Constitution in Chinese Medicine Questionnaire,designed by WANG Qi(English version) to survey TCM constitutions of the American and Canadian Caucasian in Beijing. China Journal of Traditional Chinese Medicine and Pharmacy 2012;

17. Lagrand WK, Visser CA, Hermens WT, et al. C-Reactive Protein as a Cardiovascular Risk Factor. Circulation 1999;100(1):96–102, doi:doi:10.1161/01.CIR.100.1.96

18. Borghi C, Piani F. Uric Acid and Risk of Cardiovascular Disease: A Question of Start and Finish. Hypertension 2021;78(5):1219–1221, doi:doi:10.1161/HYPERTENSIONAHA.121.17631

19. Hasan S, Naugler C, Decker J, et al. Laboratory reporting of framingham risk score increases statin prescriptions in at-risk patients. Clin Biochem 2021, doi:10.1016/j.clinbiochem.2021.06.004

20. Backus BE, Gerretsen BM. The contemporary significance of Framingham risk factors. Eur J Emerg Med 2021;28(3):167–168, doi:10.1097/MEJ.0000000000000824

21. Liu S, Liu Q. Personalized magnesium intervention to improve vitamin D metabolism: applying a systems approach for precision nutrition in large randomized trials of diverse populations. Am J Clin Nutr 2018;108(6):1159–1161, doi:10.1093/ajcn/nqy294

22. Dai Q, Zhu X, Manson JE, et al. Magnesium status and supplementation influence vitamin D status and metabolism: results from a randomized trial. Am J Clin Nutr 2018;108(6):1249–1258, doi:10.1093/ajcn/nqy274

23. Sun Y, Liu P, Zhao Y, et al. Characteristics of TCM constitutions of adult Chinese women in Hong Kong and identification of related influencing factors: a cross-sectional survey. J Transl Med 2014;12(140, doi:10.1186/1479-5876-12-140

24. Wilson PW, D’Agostino RB, Levy D, et al. Prediction of coronary heart disease using risk factor categories. Circulation 1998;97(18):1837–47, doi:10.1161/01.cir.97.18.1837

25. D’Agostino RB, Sr., Vasan RS, Pencina MJ, et al. General cardiovascular risk profile for use in primary care: the Framingham Heart Study. Circulation 2008;117(6):743–53, doi:10.1161/CIRCULATIONAHA.107.699579

26. Li L, Yao H, Wang J, et al. The Role of Chinese Medicine in Health Maintenance and Disease Prevention: Application of Constitution Theory. Am J Chin Med 2019;47(3):495–506, doi:10.1142/S0192415X19500253

27. Zhu YB, Wang Q, Chen KF, et al. [Stratified analysis of the relationship between traditional Chinese medicine constitutional types and health status in the general population based on data of 8,448 cases]. Zhong Xi Yi Jie He Xue Bao 2011;9(4):382–9, doi:10.3736/jcim20110406

28. Luo H, Li L, Li T, et al. Association between metabolic syndrome and body constitution of traditional Chinese medicine: A systematic review and meta-analysis. Journal of Traditional Chinese Medical Sciences 2020;7(4):355–365, doi:https://doi.org/10.1016/j.jtcms.2020.10.004

29. Li M, Mo S, Lv Y, et al. A Study of Traditional Chinese Medicine Body Constitution Associated with Overweight, Obesity, and Underweight. Evid Based Complement Alternat Med 2017;2017(7361896, doi:10.1155/2017/7361896

30. (NCD-RisC) NRFC. Trends in adult body-mass index in 200 countries from 1975 to 2014: a pooled analysis of 1698 population-based measurement studies with 19·2 million participants. Lancet 2016;387(10026):1377–1396, doi:10.1016/s0140-6736(16)30054-x

31. Lu Y, Wang P, Zhou T, et al. Comparison of Prevalence, Awareness, Treatment, and Control of Cardiovascular Risk Factors in China and the United States. J Am Heart Assoc 2018;7(3), doi:10.1161/JAHA.117.007462

32. Wang YC, Wei LJ, Liu JT, et al. Comparison of Cancer Incidence between China and the USA. Cancer Biol Med 2012;9(2):128–32, doi:10.3969/j.issn.2095-3941.2012.02.009

33. Rahimi-Sakak F, Maroofi M, Rahmani J, et al. Serum uric acid and risk of cardiovascular mortality: a systematic review and dose-response meta-analysis of cohort studies of over a million participants. BMC Cardiovascular Disorders 2019;19(1):218, doi:10.1186/s12872-019-1215-z

34. Xu F, Wu G, Miao J, et al. Coronary arterial disease correlates with constitutions of Traditional Chinese Medicine: A cross-sectional study in a Chinese cohort. Traditional Medicine and Modern Medicine 2018;01(03):199–205, doi:10.1142/s2575900018500118

