## Supplemental Marterial for "Associations between Traditional Chinese Medicine Body Constitution and Cardiovascular Disease Risk in a White population"

### ^4^Atrium Health, Center for Outcome Research and Evaluation, Charlotte, NC, USA

^5^ Department of Medicine, Vanderbilt University School of Medicine, Vanderbilt University Medical Center, Nashville, TN, USA

### ^6^Saint Joseph Health System, South Bend, IN, USA

^7^Department of Medicine, Division of Geriatric Medicine; Vanderbilt University Medical Center, Nashville, TN, USA

^8^Department of Medicine, Division of Gastroenterology, Hepatology, and Nutrition, Vanderbilt University Medical Center, Nashville, TN, USA

^9^Department of Biostatistics, Vanderbilt University School of Medicine, Vanderbilt University Medical Center, Nashville, TN, USA

^#^ Indicates shared first authorship.

**Personalized Prevention of Colorectal Cancer Trial** (PPCCT)

The PPCCT is a double-blind 2×2 factorial randomized controlled trial conducted at Vanderbilt University Medical Center, Nashville, TN.

**Inclusion criteria**: Aged 40 to 85 participants who had history of adenomas or hyperplastic polyps diagnosed from 1998 to 2014 or with a high risk of colorectal cancer. All participants had a calcium intake ≥ 700 mg/day and < 2000 mg/day and the calcium:magnesium intake ratio was greater than 2.6 based on two 24-hour dietary recalls.

**Exclusion criteria**: Those with a history of colectomy, inflammatory bowel disease, any organ transplantation, cancer other than non-melanoma skin cancer, gastric bypass, chronic renal diseases and hepatic cirrhosis, chronic ischemic heart disease, diarrhea, type I diabetes mellitus, pituitary dwarfism; current use of lithium carbonate therapy, blood anticoagulant drugs, digoxin and licorice; no contact information and informed consent were excluded from the study.

**Biomarker assay**

Blood samples were collected from forearm IV access site at each study visit after participants had fasted for at least 8 hours. Study visits occurred at baseline, approximately 6 weeks, and approximately 12 weeks after baseline. The blood was clotted and immediately centrifuged to separate serum, which was rapidly cooled and frozen at -80^o^C before biochemistry analysis. Serum samples were assayed for a lipid profile (low-density lipoproteins cholesterol (LDL-C), high-density lipoproteins cholesterol (HDL-C), total cholesterol (TC), and triglycerides), c-reactive protein (CRP), and uric acid at the Vanderbilt Lipid Laboratory which is standardized by the Centers for Disease Control and Prevention for lipid analysis [1]. Total cholesterol, HDL-c, triglycerides, and uric acid were measured using the Reagents of ACE® Cholesterol (#SA1010), HDL-c (#SA1038), triglycerides (#SA1023), and uric acid (#SA1025) respectively [1]. The detailed assay procedure can be referred to the appropriate Alfa Wassermann Diagnostic Technologies, LLC Clinical Chemistry Systems Operator's Manual. When triglycerides were less than or equal to 400 mg/dl, the LDL-c was calculated by subtracting HDL-c and one-fifth of triglycerides from the total cholesterol. Otherwise, the direct LDL-c was directly measured using the ACE LDL-c Reagent (#SA1040)[1]. Based on values measured from quality control samples, intra-assay and inter-assay variation coefficients were 1.28% and 4.03% for LDL-c; 0.63% and 2.19% for HDL-c; 0.67% and 5.12% for triglycerides; 0.75% and 2.06% for total cholesterol, 1.72% and 7.64% for uric acid.

Serum C-Reactive Protein (CRP) concentrations were determined by the latex particle enhanced immunoturbidimetric assay [2-4] (Pointe Scientific, Inc, Canton, MI) at the Vanderbilt Lipid Laboratory. The assay was standardized by the Centers for Disease Control and Prevention for lipid analysis [5]. Within-, and day-to-day CVs ranged from 1.16% to 5.74%, 0.8% to 5.5%, and 1.23% to 6.97%, respectively.

Serum creatinine was measured using a kinetic alkaline picrate method using a Cobas Mira Plus clinical autoanalyzer and a kit from Randox Laboratories (Crumlin, UK); CVs were less than 6%. Serum creatinine-based estimated glomerular filtration rate (eGFR) was obtained by using the modified 4-variable Modification of Diet in Renal Disease (MDRD) Study equation [6].

**General cardiovascular risk score**

General cardiovascular risk score (GCRS) were calculated based on the six coronary risk factors including age, gender, TC, HDL-C, systolic blood pressure, and smoking habits. The cutoffs for calculating GCRS were as follows: TC < 160, 160–199, 200–239, 240–279, and ≥ 280 mg/dL; HDL-C: < 40, 40–49, 50–59, and ≥ 60 mg/dL; systolic blood pressure: < 120, 120–129, 130–139, 140–159, and ≥ 160 mmHg [7]

1. Lee SA, Wen W, Xiang YB, et al. Stability and reliability of plasma level of lipid biomarkers and their correlation with dietary fat intake. Dis Markers 2008;24(2):73-9 doi: 10.1155/2008/347817[published Online First: Epub Date]|.

2. Lizana J, Hellsing K. Manual immunoephelometric assay of proteins, with use of polymer enhancement. *Clin Chem* 1974;20:1181-1186.

3. Otsuji S, Shibata H, Umeda M. Turbidimetric immunoassay of serum C-reactive protein. *Clin Chem* 1982;28:2121-2124.

4.	Malkus H, Buschbaum P, Castro A. An automated turbidimetric rate method for immunoglobulin assays. *Clin Chim Acta* 1978;88:523-530.

5. Lee SA, Wen W, Xiang YB et al. Stability and reliability of plasma level of lipid biomarkers and their correlation with dietary fat intake. *Dis Markers* 2008;24:73-79.

6. You L, Zhu X, Shrubsole MJ, et al. Renal function, bisphenol A, and alkylphenols: results from the National Health and Nutrition Examination Survey (NHANES 2003-2006). Environ Health Perspect 2011;119(4):527-33 doi: 10.1289/ehp.1002572[published Online First: Epub Date]|.

7. Flueckiger P, Longstreth W, Herrington D, Yeboah J. Revised Framingham Stroke Risk Score, Nontraditional Risk Markers, and Incident Stroke in a Multiethnic Cohort. Stroke 2018;49(2):363-69 doi: 10.1161/STROKEAHA.117.018928[published Online First: Epub Date]|.

| Supplemental Table 1 Distribution of TCM body constitution, stratified by age and gender | | | | |  |  |  |  |
| --- | --- | --- | --- | --- | --- | --- | --- | --- |
| Characteristic | American in US | | | | American/Canadian in China | | | |
|  | <50 y | ≥50 y | Man | Women | <50 y | ≥50 y | Man | Women |
| Sample size, n | 13 | 178 | 100 | 91 | 322 | 74 | 156 | 240 |
| TCM Constitution type, % |  |  |  |  |  |  |  |  |
| *Gentleness* | 15.4 | 30.9 | 42.0 | 16.5 | 50.9 | 51.4 | 62.8 | 43.4 |
| *Qi-deficiency* | 0.0 | 14.6 | 16.0 | 11.0 | 9.9 | 8.1 | 9.0 | 10.0 |
| *Yang-deficiency* | 7.7 | 4.5 | 1.0 | 8.8 | 13.7 | 13.5 | 9.0 | 16.7 |
| *Yin-deficiency* | 0.0 | 6.7 | 2.0 | 11.0 | 5.0 | 2.7 | 2.6 | 5.8 |
| *Phlegm-Dampness* | 23.1 | 7.3 | 6.0 | 11.0 | 3.1 | 13.5 | 5.1 | 5.0 |
| *Dampness-Heat* | 0.0 | 3.4 | 5.0 | 1.1 | 4.3 | 0.0 | 2.6 | 4.2 |
| *Blood-Stasis* | 38.5 | 15.7 | 8.0 | 27.5 | 2.5 | 2.7 | 1.3 | 3.3 |
| *Qi-depression* | 0.0 | 6.7 | 8.0 | 4.4 | 3.7 | 5.4 | 3.8 | 4.2 |
| *Special Diathesis* | 15.4 | 10.1 | 12.0 | 8.8 | 6.8 | 2.7 | 3.8 | 7.5 |

Continuous variables were presented as Mean ± SD; categorical variables were presented as %

TCM: Traditional Chinese medicine

| Supplemental Table 2 Spearman item-scale correlation and scaling success rate of the TCM | | |
| --- | --- | --- |
| Body constitution classification by TCM | Baseline (n=191) | |
|  | Item-scale correlation | Scaling success rate (%) |
| Gentleness | 0.510-0.658 | 75 |
| Qi-deficiency | 0.342-0.640 | 100 |
| Yang-deficiency* | 0.155-0.785 | 87.5 |
| Yin-deficiency | 0.409-0.682 | 100 |
| Phlegm-wetness | 0.498-0.682 | 100 |
| Wetness-heat | 0.400-0.728 | 100 |
| Blood-stasis | 0.497-0.720 | 87.5 |
| Qi-depression | 0.500-0.822 | 100 |
| Special diathesis | 0.442-0.730 | 100 |
| TCM: Traditional Chinese medicine |  |  |

| Supplemental Table 3 The Intra-class correlation (ICC) coefficients of the TCM | |
| --- | --- |
| Body constitution classification by TCM | ICC (95% CI) |
|  | n=164 |
| Gentleness | 0.77 (0.70-0.82 |
| Qi-deficiency | 0.74 (0.66-0.80) |
| Yang-deficiency | 0.70 (0.61-0.77) |
| Yin-deficiency | 0.74 (0.66-0.80) |
| Phlegm-wetness | 0.80 (0.74-0.85) |
| Wetness-heat | 0.75 (0.67-0.81) |
| Blood-stasis | 0.77 (0.70-0.82) |
| Qi-depression | 0.80 (0.74-0.85) |
| Special diathesis | 0.71 (0.62-0.78) |
| TCM: Traditional Chinese medicine | |
